# Early-onset schizophrenia is associated with immune-related rare variants in a Chinese sample

**DOI:** 10.1101/2023.11.21.23298115

**Authors:** Yuanxin Zhong, Justin D. Tubbs, Perry B.M. Leung, Na Zhan, Tomy C.K. Hui, Karen K. Y. Ho, Karen S. Y. Hung, Eric F. C. Cheung, Hon-Cheong So, Simon S.Y. Lui, Pak C. Sham

**Affiliations:** Department of Psychiatry, School of Clinical Medicine, LKS Faculty of Medicine, The University of Hong Kong, Hong Kong SAR; Psychiatric and Neurodevelopmental Genetics Unit, Center for Genomic Medicine, Massachusetts General Hospital, Boston, MA, USA; Department of Psychiatry, Harvard Medical School, Boston, MA, USA; Stanley Center for Psychiatric Research, Broad Institute of MIT and Harvard, Cambridge, MA, USA; Department of Psychosis Studies, Institute of Psychiatry, Psychology and Neuroscience, King’s College London, London, UK; School of Biomedical Sciences, The Chinese University of Hong Kong, Shatin, Hong Kong SAR; Department of General Adult Psychiatry, Castle Peak Hospital, Hong Kong SAR; Department of Psychiatry, The Chinese University of Hong Kong, Shatin, Hong Kong SAR; Centre for PanorOmic Sciences, Li Ka Shing Faculty of Medicine, The University of Hong Kong, Hong Kong SAR; State Key Laboratory of Brain and Cognitive Sciences, The University of Hong Kong, Hong Kong SAR

**Keywords:** schizophrenia, exome-sequencing, age at onset, immune response

## Abstract

**Background:** Rare variants are likely to contribute to schizophrenia (SCZ), given the large discrepancy between the heritability estimated from twin and GWAS studies. Furthermore, the nature of the rare-variant contribution to SCZ may vary with the “age-at-onset” (AAO), since early-onset has been suggested as being indicative of neurodevelopment deviance.

**Objective:** To examine the association of rare deleterious coding variants in early- and adult-onset SCZ in a Chinese sample.

**Method:** Exome sequencing was performed on DNA from 197 patients with SCZ spectrum disorder and 82 healthy controls (HC) of Chinese ancestry recruited in Hong Kong. We also gathered AAO information in the majority of SCZ samples. Patients were classified into early-onset (EOS, AAO<18) and adult-onset (AOS, AAO>18). We collapsed the rare variants to improve statistical power and examined the overall association of rare variants in SCZ versus HC, EOS versus HC, and AOS versus HC at the gene and gene-set levels by Sequence Kernel Association Test. The quantitative rare-variant association test of AAO was also conducted. We focused on variants which were predicted to have a medium or high impact on the protein-encoding process as defined by Ensembl. We applied a 100000-time permutation test to obtain empirical p-values, with significance threshold set at p < 1e^-3^ to control family-wise error rates. Moreover, we compared the burden of targeted rare variants in significant risk genes and gene sets in cases and controls.

**Results:** Based on several binary-trait association tests (i.e., SCZ vs HC, EOS vs HC and AOS vs HC), we identified 7 candidate risk genes and 20 gene ontology biological processes (GOBP) terms, which exhibited higher burdens in SCZ than in controls. Based on quantitative rare-variant association tests, we found that alterations in 5 candidate risk genes and 7 GOBP pathways were significantly correlated with AAO. Based on biological and functional profiles of the candidate risk genes and gene sets, our findings suggested that, in addition to the involvement of perturbations in neural systems in SCZ in general, altered immune responses may be specifically implicated in EOS.

**Conclusion:** Disrupted immune responses may exacerbate abnormal perturbations during neurodevelopment and trigger the early onset of SCZ. We provided evidence of rare variants increasing SCZ risk in the Chinese population.

## Introduction

Schizophrenia (SCZ) is a severe mental disorder affecting around 0.7-1% of the general population worldwide, and is associated with sufferings and increased burden on patients, their caregivers and society (WHO, 2019). SCZ is highly familial, with heritability estimates of 60–80% (Cannon, Kaprio, Lönnqvist, Huttunen, & Koskenvuo, 1998; Lichtenstein et al., 2009). The identification of the causal genes for SCZ has the potential to unveil the putative pathophysiological mechanisms of SCZ.

Genome-wide association study (GWAS) has been successfully used to investigate the genetic component of SCZ vulnerability (Kotlar, Mercer, Zwick, & Mulle, 2015). Although more than 200 SCZ-related genetic loci have been identified (Trubetskoy et al., 2022), the SNP-based heritability in samples with European ancestry was estimated at the modest value of 0.24. Moreover, the polygenic score (PGS) on the “all-ancestry” primary SCZ GWAS only explained a median of 0.024 of variance of liability, indicating the presence of “missing heritability” in SCZ. Relatively small effects of common risk variants and genes among the populations have been reported (Purcell et al., 2009), which may limit their translational potentials. Whole-exome sequencing (WES) can identify rare variants which are not detected by GWAS but likely have a larger effect size than common variants, and therefore has the potential to promote precision medicine (Senormanci, Karakas Celik, Valipour, Dogan, & Senormanci, 2018). A recent large-scale whole-exome meta-analysis with 24,248 SCZ cases and 97,322 controls identified ten genes that carry rare coding variants conferring risk for SCZ (Singh et al., 2022).

The identification of the genetic architecture of SCZ has been complicated by the problem of phenotypic heterogeneity (Jablensky, 2006). Although the DSM-5 classification system has obliterated the subtyping of SCZ based on psychopathological symptoms, the current empirical evidence supports that SCZ patients can differ substantially in the illness course (Dickinson et al., 2018), cognitive profiles (Lim et al., 2021) and brain structures (Pan et al., 2020; Shi et al., 2023). Dissecting SCZ into more homogeneous subtypes might be beneficial for clarifying the genetic component of SCZ vulnerability.

The age at onset (AAO) in SCZ is an important feature for dissecting the phenotype heterogeneity in SCZ (Chen, Selvendra, Stewart, & Castle, 2018; Zhan, Sham, So, & Lui, 2023). For instance, although the typical onset age for SCZ is 21-25 years for men and 25-30 for women, some patients can develop SCZ during childhood and adolescence, i.e., patients with early-onset SCZ (EOS) (with AAO <18) (Werry, 1992). The SCZ subtypes of EOS and adult-onset SCZ (AOS; AAO >=19) have substantial phenotypic differences (Howard et al., 2018). For instance, patients with EOS have more severe symptoms (Kendhari, Shankar, & Young-Walker, 2016), cognitive dysfunctions (Frangou, 2010), lower educational attainment (Coulon et al., 2020), more premorbid language problems (Hollis, 1995) and other neurodevelopmental abnormalities (Vourdas, Pipe, Corrigall, & Frangou, 2003), than patients with AOS. Besides, relative to AOS, patients with EOS have higher chance to evolve to treatment-resistant SCZ (Iasevoli et al., 2022). Importantly, family studies reported a higher familial risk for childhood- and adolescent-onset SCZ than for adult-onset SCZ (Nicolson et al., 2003). Therefore, it can be hypothesized that EOS may have a greater burden of rare deleterious genetic variants, and that these variants are most likely to disrupt brain development than those associated with AOS (Birnbaum & Weinberger, 2017).

The genetic architecture of AAO in SCZ remains under-studied. Different study designs and methods were applied to detect the genetic basis of AAO (Zhan et al., 2023). For instance, previous candidate-gene hypothesis-driven studies examined the associations between AAO and SCZ candidate genes which involved a wide variety of functions, including dopamine receptors (e.g., *DRD2*, *DRD3*) (Alfimova, Nikitina, Lezheiko, Simashkova, & Golimbet, 2023; Michalczyk et al., 2020), neurotransmitter metabolism (e.g., *COMT*, *DBH*) (Barlas et al., 2012; Numata et al., 2007; Pelayo-Terán et al., 2010; Tylec, Jeleniewicz, Mortimer, Bednarska-Makaruk, & Kucharska, 2017), cell-cell signaling mediation (e.g., *NRG1*) (Papiol et al., 2011; Weickert, Tiwari, Schofield, Mowry, & Fullerton, 2012; Yoshimi et al., 2016), and immune processes (e.g., *TNF-RII*) (Wassink, Crowe, & Andreasen, 2000). However, such candidate-gene approach had yielded low replicability and inconsistent findings. To-date, only five GWAS on SCZ AAO have been published (Bergen et al., 2014; Guo et al., 2021; Sada-Fuente et al., 2023; Wang, Liu, Zhang, Aragam, & Pan, 2011; Woolston et al., 2017). The first GWAS, based on 1162 European-American individuals, reported 104 SNPs at a suggestive p-value threshold (p = 1e^-4^) but failed to replicate the findings in an independent sample (Wang et al., 2011). A recent study conducted the GWAS of SCZ OAA in a larger sample from European ancestry (N = 4740), but no genome-wide significant (GWS) locus was identified (Sada-Fuente et al., 2023). The first GWAS attempt on SCZ AAO in the East-Asian population utilized a small sample (N = 185). This study identified 14 SNPs at a suggestive p-value threshold (p = 1e^-4^) and found that the earlier onset group had a higher genetic risk score, calculated based on the 14 SNPs (Woolston et al., 2017). A two-stage GWAS of EOS in Han Chinese population was conducted and four GWS loci were identified in the combined samples (2159 EOS cases and 6561 controls). The *MTHFR*, *TDGF1*, *ANGPTL2*, and *RALGPS1* genes, which are the four eQTL targets of the GWS risk variants, are crucial for brain development. This study provides valuable insight into the genetic basis of EOS (Guo et al., 2021); however, more research is necessary.

To our knowledge, few rare variant studies on SCZ AAO (Alkelai et al., 2023; Cohen, Singh, Öngür, Konstantin, & Gardner, 2022; Lee et al., 2012; Martin, Robinson, Reutens, & Mowry, 2014) have been conducted. One study reported that loss-of-function variants in the *SCHEMA* gene were associated with phenotypic features of SCZ, such as earlier AAO and poorer treatment outcomes (Cohen et al., 2022). Another study performed exome sequencing (ES) on 37 Israeli Jewish families with a proband diagnosed with childhood-onset schizophrenia, and found significant variants involving the *ANKRD11*, *GRIA2*, *CHD2*, *CLCN3*, *CLTC*, *IGF1R*, and *MICU1* genes (Alkelai et al., 2023).

The aforementioned evidence gathered using GWAS, ES, or WES were largely based on samples with European ancestry. The association of SCZ AAO with rare variants in samples with Chinese ancestry has not yet been examined. Moreover, genetic studies on AAO in SCZ provided some evidence supporting that EOS and AOS have shared variants (Ahn, An, Shugart, & Rapoport, 2016; Asarnow & Forsyth, 2013; Chaumette et al., 2020; Forsyth & Asarnow, 2020; Guo et al., 2021; Yadav, Kumar, Gupta, & Rai, 2016) such as *MTHFR* (Guo et al., 2021; Yadav et al., 2016), but direct comparison between EOS and AOS had seldom been studied. Taken together, several issues remain unresolved. First, the rare coding risk genes and pathways associated with AAO in SCZ remains elusive. Second, the rare variants in SCZ patients with Chinese ancestry were under-studied. Third, it remained unclear whether EOS patients and AOS patients would have different candidate risk genes or pathways with an increased burden of rare variants, because previous studies in this area adopted a family design and did not make direct comparison between EOS and AOS patients.

This study aimed to examine the association of rare variants with predicted damaging impacts in the whole exome between SCZ patients and healthy controls (HC) with Chinese ancestry. Using AAO to categorize the SCZ sample into EOS and AOS groups, we aimed to examine the candidate risk genes and pathways (gene-sets) bearing increased burden of rare variants contributing to EOS and AOS respectively. Furthermore, we aimed to examine the rare-variant associations of AAO (as a continuous variable), for understanding the potential genetic factors causing deviated neurodevelopment at different stages in SCZ.

## Materials and methods

### 1. Participants

Our sample comprised 197 SCZ patients recruited from a research-orientated clinical programme at Castle Peak Hospital Hong Kong (Lui, Sham, Chan, & Cheung, 2011) and 82 healthy controls in the neighbouring community. The DSM-V (American Psychiatric Association & Association, 2013) diagnosis of SCZ in clinical participants was ascertained by two experienced psychiatrists using a structured interview, supplemented by medical records. The exclusion criteria for SCZ participants included (1) history of illicit substance use in the past 12 months, (2) electroconvulsive therapy (ECT) in the past 6 months, (3) neurological disorders, (4) brain injury with >30 min of unconsciousness, and (5) mental retardation. Healthy controls (HC) were interviewed by qualified psychiatrists to ensure the absence of personal and family history of SCZ and related psychosis. HC also completed the Schizotypal Personality Questionnaire (SPQ) (Raine, 1991) and they all scored < 45. Demographics and clinical variables, including AAO, were gathered from medical records. Based on the cut-off AAO of 18, we categorized the SCZ sample into childhood/adolescent-onset schizophrenia (EOS, n = 47) and adult-onset schizophrenia (AOS, n = 125) subgroups.

This study was approved by the New Territories West Research Ethics Committee (Protocol number: NTWC/CREC/823/10; NTWC/CREC/1293/14; UW14-325). All participants provided written informed consent. No monetary incentive was provided to the participants. The flow chart in **Figure 1** illustrates the design of this study.

**Figure 1.**
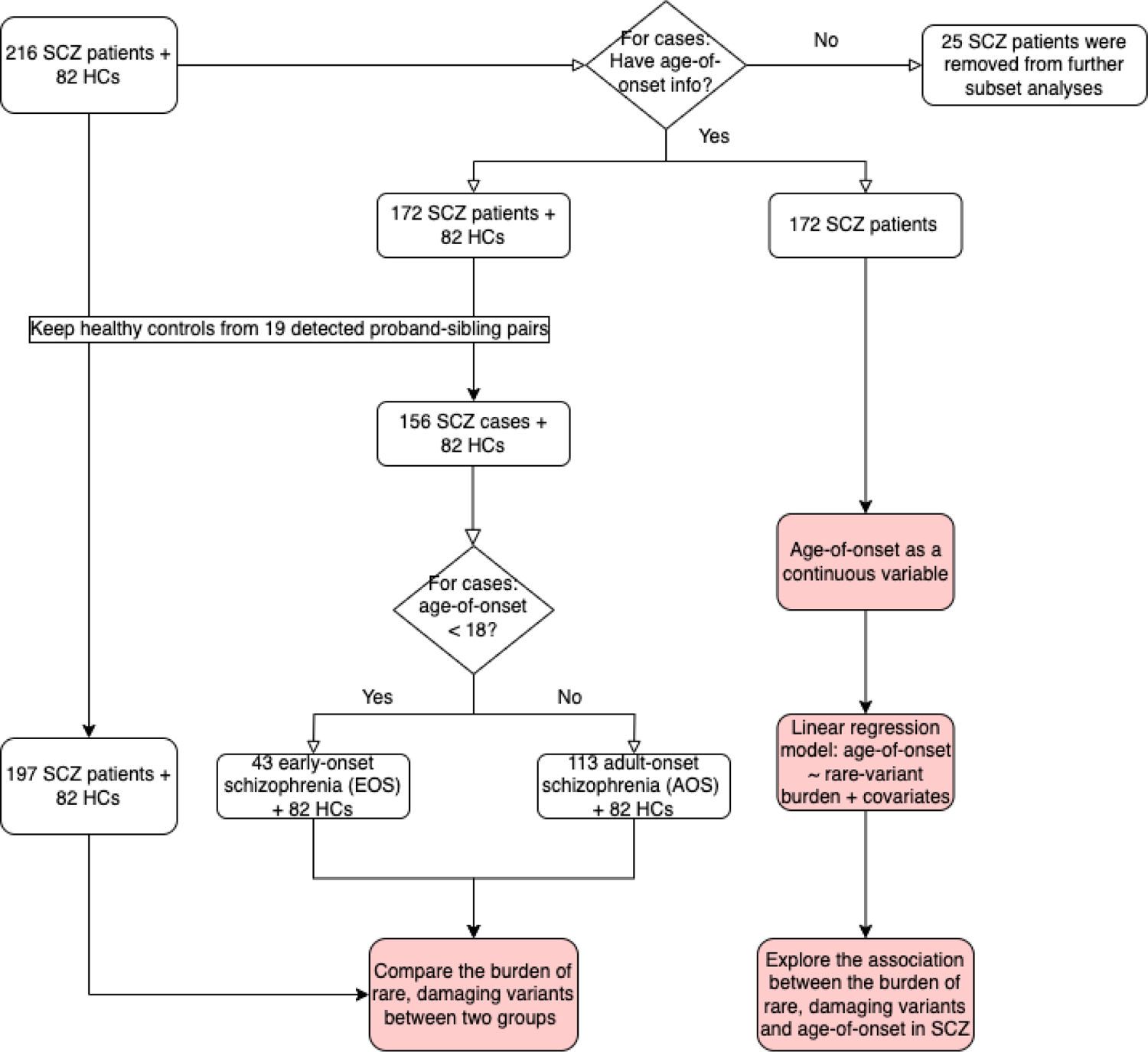
The pipeline diagram presents four different combinations of samples utilized in the corresponding analyses in the current study. SCZ, schizophrenia; HC, healthy control.

### 2. Exome-sequencing and data processing

Exonic DNA was captured using the IDT x Gen Exome Research Panel v1.0 including supplemental probes and sequenced using Illumina HiSeq sequencing platform. Read files (Fastq) were generated subsequently. Firstly, we applied FastQC to check the quality of DNA sample, focusing on base quality score, GC content, N content, and sequence duplication level. Burrows-Wheeler Aligner (BWA) package version 0.7.12 (Li & Durbin, 2010) was performed to align sequence reads to the human reference genome (hg19). Picard (https://broadinstitute.github.io/picard/) was used to identify duplicate reads to filter out those that were probably caused by PCR bias. With Samtools, generated BAM files were further modified (Li et al., 2009). To improve the quality of variants calls, we utilized the base quality score recalibration (BQSR) function, implemented in the Genome Analysis Toolkit (GATK) (McKenna et al., 2010), for detecting artificial errors caused by the sequencing machine and recalibrating base quality (Phred scale) scores. We also applied the Picard algorithm to coordinate, sort, and index the curated BAM files. The variant calling was processed based on the GATK Best Practices recommendations (McKenna et al., 2010). We used GATK to calculate variant quality score recalibration (VQSR) and plink for other measures to evaluate the quality of variants and detect potential contamination. We chose 95% as the sensitivity threshold. Genotypes with read depth (DP) < 8 or genotype quality (GQ) < 20 were eliminated. Filters of allele balance (AB >0.25) and missingness rate of > 0.1 were also applied to exclude the low-quality variants. Samples with a genotyping rate < 0.95 were removed.

The qualified variants were annotated by the Ensembl VEP (McLaren et al., 2016). Rare variants were defined as those having a MAF < 0.05 in any of the selected databases in the 1000 Genomes Project and gnomAD (all populations and the East Asian population). We defined the deleterious variants as variants rated as “MODERATE” or “HIGH” impact based on the IMPACT rating estimated by Ensembl (https://asia.ensembl.org/info/genome/variation/prediction/predicted_data.html).

### 3. Gene-based and Gene-set-based association test

Collapsing rare variants can improve the statistical power of finding disease-related genes with exome sequencing data (Sun, Sung, Tintle, & Ziegler, 2011). We applied an annotated gene as the testing unit. The genes with at least three rare mutant alleles (deleterious variants as defined above) in the entire sample were included for association testing using Rvtests (Zhan, Hu, Li, Abecasis, & Liu, 2016) (see **Figure 1**). Sequencing kernel association test (SKAT) was used to evaluate the association between genes and SCZ risk. We used a 100000-time permutation test to obtain empirical p-values for the SKAT tests. The significance level was set at 1e^-3^ (Purcell et al., 2014).

We used all GO (gene ontology) biological pathway (BP) terms (Liberzon et al., 2011) for the gene-set-based association tests. We included the gene sets with at least three rare damaging alleles across the samples. SKAT and 100000-time permutation test were used for the analyses. Moreover, we compared the burden of targeted rare variants in significant risk genes and gene sets in cases and controls. We focused on candidate genes and pathways with a higher burden of rare variants in cases than controls, which is commonly associated with a decrease in gene expression and thus exerts a risk-increasing effect on disease (Rivas et al., 2015).

To gain deeper insight into the genetic architecture of SCZ, we conducted analyses of four different combinations of sample (see **Figure 1**), (1) SCZ patients (n = 197) versus HC (n = 82); (2) EOS patients (n = 43) versus HC (n = 82); (3) AOS patients (n = 113) versus HC (n = 82); and (4) EOS + AOS (n = 172). We used AAO as a continuous variable and examined its correlation with the rare variants at gene and gene-set based levels.

### 4. The association of candidate risk genes with other traits

The phenotype-wide association analysis (PheWAS) was performed to look for GWAS signals in other traits. Candidate risk genes were used to search the GWAS Catalog (Sollis et al., 2022), PheWAS Atlas (Watanabe et al., 2019), and PhenoScanner v2 (Kamat et al., 2019) for associations (P < 5e^-8^) with other 4756 traits and diseases across different systems.

## Results

### 1. Data description

After quality control, 216 SCZ patients and 82 HCs were retained in the sample. AAO information was available on 172 SCZ patients. Relationship checks on the genotype data revealed 19 proband-sibling pairs. To retain the statistical power, we kept HC data from the sibling pairs. This resulted in 197 SCZ patients (45.2% female; mean age, 26.7 years) and 82 HCs (48.8% female; mean age, 27.3 years) with 128,192 rare variants remaining in the final sample (See Figure 1). A total of 46,520 rare variants (MAF < 0.05) were annotated, including 72 start-loss variants, 30 stop-loss variants, 268 stop-gain variants, 23491 missense variants, and 22659 synonymous variants. Benign coding variants (22659 synonymous variants) were excluded from the subsequent association tests.

### 2. Gene-based association test results

Gene-based analysis of 8,077 rare, deleterious variants were conducted in four different combinations of sample (see **Figure 1**) and showed the following results. As we compared SCZ (n = 197) and HC (n = 82), qualifying variants map to 4314 genes. *NUF2* (P_FWER_ = 1e^-5^), *PACRGL* (P_FWER_ = 8.8e^-4^), and *MSH4* (P_FWER_ = 9.2e^-4^) showed significant associations with SCZ (see Table 1), yet the burden is higher in HC than in SCZ, suggesting a “protective” effect. As we compared EOS (n = 43) and HC (n = 82), variants map to 3123 genes. The following six genes achieved significance after permutation: *PADI2* (P_FWER_ = 3.2e^-4^), *RSPH9* (P_FWER_ = 4.1e^-4^), *SYNM* (P_FWER_ = 4.8e^-4^), *CD4* (P_FWER_ = 5.5e^-4^), *ANKS3* (P_FWER_ = 6.55e^-4^), and *USP10* (P_FWER_ = 9.9e^-4^). These genes showed a higher burden in SCZ than in HC, suggesting that they would increase SCZ risk. As we compared AOS (n = 113) and HC (n = 82), 3,736 genes were included. *TPCN2* (P_FWER_ = 4.1e^-4^) was a strong candidate gene involved in SCZ. As we examined the association between the rare variants in genes and AAO in the combined sample of EOS and AOS patients (n = 172), five genes were related to AAO, i.e., *STOX1* (P_FWER_ = 3.6e^-4^), *WDHD1* (P_FWER_ = 5.5e^-4^), *ZDHHC14* (P_FWER_ = 6.9e^-4^), *CHRM2* (P_FWER_ = 7.4e^-4^), *MTHFR* (P_FWER_ = 7.7e^-4^). Among these five genes, *STOX1* showed the strongest signal which survived Bonferroni corrections (P-value threshold = 0.05/3508 = 1.43e^-5^) in the CMC burden test (P_unadj_ = 4.67e^-6^). The results are shown in **Table 1** and **Figure 2**.

**Figure 2.**
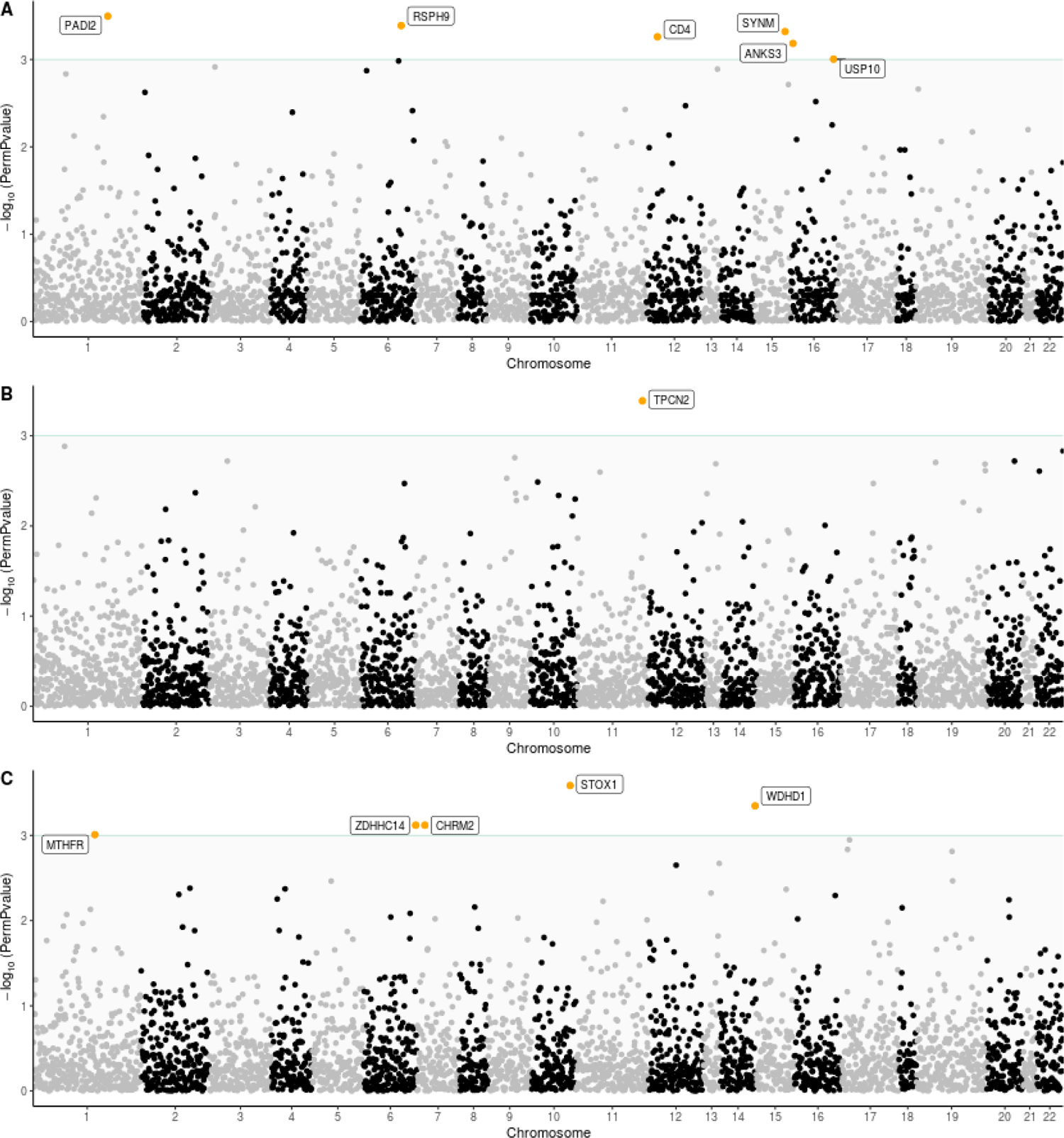
Exome-wide gene-based association tests. (A) Early-onset schizophrenia vs Healthy controls; (B) Adult-onset schizophrenia vs Healthy controls; (C) Age at onset (AAO in schizophrenia cases as a continuous variable). The genes highlighted were the candidate risk genes significantly correlated with the phenotypes (P_FWER_ < 0.001). The horizontal line indicates a significant level of 0.001.

**Table 1.** Gene-based rare-variant burden comparison.

| Gene | Variants | Cases | Controls | P<sub>FWER</sub> |
| --- | --- | --- | --- | --- |
| **(1) SCZ patients (N = 197) verse HCs (N = 82)** |  |  |  |  |
| *NUF2* | 1:163313539 | 4 (2.03%) | 13 (15.85%) | 1.00E-05 |
| *PACRGL* | 4:20706388, 4:20706397,<br>4:20714522 | 7 (3.55%) | 11 (13.41%) | 0.00088 |
| *MSH4* | 1:76269460, 1:76378502 | 0 (0.00%) | 3 (3.66%) | 0.00092 |
| **(2) EOS (N = 43) verse HCs (N = 82)** |  |  |  |  |
| *PADI2* | 1:17395724, 1:17413127,<br>1:17420122, 1:17431434 | 7 (16.28%) | 2 (2.44%) | 0.00032 |
| *RSPH9* | 6:43623326, 6:43623417 | 6 (13.95%) | 0 (0.00%) | 0.00041 |
| *SYNM* | 15:99666979 | 7 (16.28%) | 0 (0.00%) | 0.00048 |
| *CD4* | 12:6925407 | 5 (11.63%) | 0 (0.00%) | 0.00055 |
| *ANKS3* | 16:4748011, 16:4755108 | 4 (9.30%) | 0 (0.00%) | 0.000655 |
| *USP10* | 16:84778697 | 37 (86.05%) | 45 (54.88%) | 0.00099 |
| **(3) AOS (N = 113) verse HCs (N = 82)** |  |  |  |  |
| *NUF2* | 1:163313539 | 3 (2.65%) | 13 (15.85%) | 0.00034 |
| *TPCN2* | 11:68854029 | 16 (14.16%) | 1 (1.22%) | 0.00041 |
| *OR1K1* | 9:125563212 | 2 (1.77%) | 10 (12.20%) | 0.00091 |
| **AAO (continuous variable)** |  | **EOS** | **AOS** | **P<sub>FWER</sub>** |
| **(4) EOS + AOS (N = 172)** |  |  |  |  |
| *STOX1* | 10:70646373 | 12 (25.5%) | 1 (0.80%) | 0.00026 |
| *WDHD1* | 14:55458038, 14:55458040,<br>14:55462410 | 10 (21.28%) | 0 (0.00%) | 0.00045 |
| *CHRM2* | 7:136700433, 7:136700603 | 10 (21.28%) | 10 (8.00%) | 0.00076 |
| *ZDHHC1*<br>4 | 6:158074592 | 7 (14.89%) | 0 (0.00%) | 0.00076 |
| *MTHFR* | 1:11850750, 1:11856343,<br>1:11863566 | 4 (8.51%) | 0 (0.00%) | 0.00098 |

The Cases and Controls columns show the number and proportion of subjects with at least one rare damaging variant in the case and control groups, respectively. SCZ, schizophrenia; HC, healthy control; EOS, Early-onset schizophrenia; AOS, Adult-onset schizophrenia; AAO, age at onset; P<sub>FWER</sub>, p values using permutation test to control family-wise error rates.

### 3. Gene-set-based association test results

Gene-based analysis of 23,861 deleterious variants were conducted in four different combinations of sample (see **Figure 1**) and showed the following pathways. As we compared SCZ (n = 197) and HC (n = 82), we found three GO BP terms achieved significance. As we compared EOS (n = 43) and HC (n = 82), 12 GO BP terms showed significant associations. Among them, the top three ranked terms are related to immune regulation (GOBP-negative regulation of response to cytokine stimulus, P_FWER_ = 9e^-5^; GOBP-negative regulation of leukocyte chemotaxis, P_FWER_ = 2.7e^-4^; GOBP-negative regulation of lymphocyte migration, P_FWER_ = 2.9e^-4^). As we compared AOS (n = 113) and HC (n = 82), five GO BP terms showed the significant signals. Among them, the GOBP-organophosphate catabolic process had the lowest p-value (P_FWER_ = 2e^-5^). As we examined the association of rare variants in gene sets with AAO in the combined sample of EOS and AOS patients (n = 172), six GOBP terms were significantly associated with AAO, including GOBP-regulation of neurotransmitter transport (P_FWER_ = 5.2e^-4^), GOBP-response to acetlcholine (P_FWER_ = 5.4e^-4^), and GOBP-response to catecholamine (P_FWER_ = 7.5e^-4^). The results are shown **Figure 3**.

**Figure 3.**
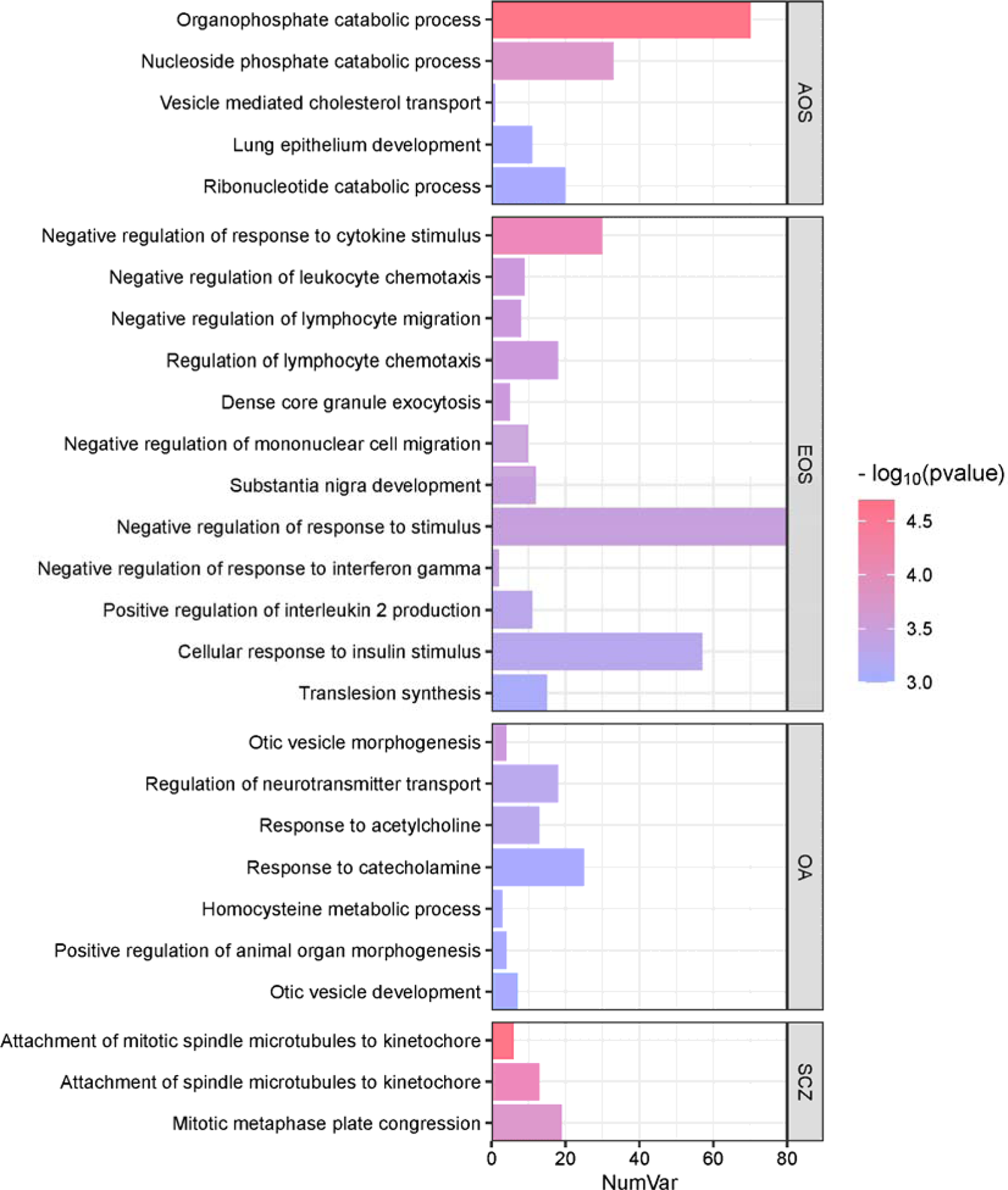
Exome-wide gene-set-based association tests. The gene sets were significantly correlated with the corresponding phenotypes (P_FWER_ < 0.001). X-axis demonstrates the number of variants included in the gene set. Within each sub-group, the gene sets are ordered by p values. AOS: Adult-onset schizophrenia vs. Healthy controls; EOS: Early-onset schizophrenia vs. Healthy controls; SCZ: SCZ (the full schizophrenia sample, n = 197) vs. HC; OA: onset of age, a continuous variable.

### 4. Co-localization of candidate risk genes with other traits

The PheWAS searched through 4756 GWAS datasets and found that *WDHD1* was significantly associated with other traits, for example, mean platelet volume, mean corpuscular volume, and resting heart rate (p < 5e^-8^). After Bonferroni correction (p-value threshold = 0.05/4756 = 1.05e^-5^), 11 traits survived and were enriched in the immunological domain (hypergeometric p = 9.13e^-5^). Furthermore, significant associations between *RSPH9* and several traits within the immunological domain were detected. Although other candidate genes were found to be related to other traits, we failed to identify specific enrichment patterns.

## Discussion

This study adds to the small number of studies on SCZ rare variants in the Chinese population. We collapsed rare pathogenic variants and conducted gene-based and gene-set-based exome-wide association analyses to identify candidate risk genes and biological pathways for SCZ. Our findings partly addressed the issue of missing heritability of SCZ and complemented other GWAS findings. In short, we identified 7 candidate risk genes and 20 potential biological pathways exhibiting a higher burden of rare damaging variants in SCZ patients. Using quantitative rare-variant association analyses, we identified 5 candidate genes and 7 GOBP pathways were particularly relevant to AAO in SCZ. These candidate genes and GOBP pathways provided useful insight for putative mechanisms which may give rise to altered neurodevelopment in SCZ. The functional profiles of our candidate genes suggested that the perturbations of neural processes are involved in the aetiology of SCZ, and abnormal immune responses may play a role in EOS.

We found three genes (*NUF2, PACRGL, MSH4*) having higher burden in HC than in SCZ, implicating a protective effect. However, a recent exome sequencing meta-analysis suggested reported that *NUF2* was nominally significant in elevating SCZ risk (odd ratio = 2.10, p = 0.051) (Singh, Neale, & Daly, 2020). In the outer kinetochore, NUF2 encodes a component of the NDC80 complex, which is crucial for proper microtubule binding and the spindle assembly checkpoint. Abnormal changes in neurite length and synaptic shape could be caused by kinetochore protein mutations (Zhao, Oztan, Ye, & Schwarz, 2019). A few reasons may explain the divergent findings regarding the direction of effects of *NUF2*. First, our small sample size increased the likelihood of sampling bias. Second, our Chinese sample differed in ancestry from the previous sample (Singh et al., 2020).

Relative to HC, EOS carried a higher rare-variant burden in 6 genes, namely *PADI2*, *RSPH9*, *SYNM*, *CD4*, *ANKS3*, and *USP10*. The gene of *PADI2* (Peptidyl Arginine Deiminase 2) encodes protein belongs to the peptidyl arginine deiminase family of enzymes, and is involved in the onset and progression of neurodegenerative disorders (Christophorou et al., 2014; Falcão et al., 2019; Xu et al., 2016; Yu et al., 2017). On the other hand, *RSPH9* (Radial Spoke Head Component 9) is involved in the motility of olfactory and neural cilia, and the structural integrity of ciliary axonemes. Impaired motile cilia may disrupt cerebrospinal fluid (CSF) circulation, and may cause ventricular dilation and hydrocephalus (Kumar et al., 2021). Some of the isoforms of *SYNM* (Synemin) are specifically expressed in the brain. Importantly, early production of synemin isoforms H/M serves as a signal for migration and differentiation of neural precursor cells (Izmiryan et al., 2010). The gene of *CD4* encodes the CD4 membrane glycoprotein of T lymphocytes, which is widely expressed in different immune components and brain regions. It may mediate secondary neuronal injury in infectious and immune-mediated diseases of the central nervous system (Buttini et al., 1998). The gene of *ANKS3* (Ankyrin Repeat And Sterile Alpha Motif Domain Containing 3) is extensively-expressed in Rod photoreceptor cells, and may be involved in visual perception, and thus the formation of certain symptoms of SCZ (e.g., visual distortions, hallucinations) (Adámek, Langová, & Horáček, 2022; Javitt, 2007). Lastly, *USP10* (ubiquitin-specific peptidase 10) constitutes part of the ubiquitin-proteasome system (UPS), which collaborates with lysosomes to break down proteins that are misfolded, damaged, or having aberrant amino acid sequences (Olanow & McNaught, 2006).

Our findings of gene-set analyses provided a consistent account on the biological and functional profiles of the candidate risk genes identified in the gene-based association tests. Together, findings of the two analyses consistently supported the involvement of the immune system in SCZ, and the potential interactions between neural and immune systems in EOS. Damaging rare variants would likely dysfunction a gene, resulting in abnormal immune responses, accumulation of cytotoxic immune proteins, and damages in the neural system. Such multi-system pathological processes may be involved in SCZ, and may even advance the first presentation of symptoms (i.e., giving rise to earlier AAO). Our findings therefore generally support the neurodevelopmental model of SCZ (Birnbaum & Weinberger, 2017; Weinberger, 2017).

Comparing AOS with HC, we found *TPCN2* significantly increased the risk of SCZ. This gene *TPCN2* (Two Pore Segment Channel 2) is expressed in dopaminergic neurons of the substantia nigra (Patel & Kilpatrick, 2018). In sporadic Parkinson disease, abnormal *TPCN2* channel conductance contributes to iron-associated cellular changes and neuronal cell death in the substantia nigra (Rivero-Ríos, Fernández, Madero-Pérez, Lozano, & Hilfiker, 2016). Abnormal dopaminergic functions have been consistently reported in SCZ (Conn, Burne, & Kesby, 2020; van Hooijdonk et al., 2023; Williams et al., 2014).

We identified 5 genes which were significantly associated with AAO in SCZ. Apart from *CHRM2*, the counts of rare damaging rare variants within each of these candidate risk genes (i.e., *STOX1*, *WDHD1*, *ZDHHC14*, and *MTHFR*) were positively correlated with AAO. *CHRM2* (Cholinergic Receptor Muscarinic 2) is expressed in inhibitory and excitatory neurons, and cardiomyocytes, and is involved in synaptic function. *CHRM2* also belongs to the muscarinic receptors family, and is involved in acetylcholinergic functions in the central and peripheral nervous system (Brown, 2010). Interestingly, *MTHFR* (Methylenetetrahydrofolate Reductase) is a gene well-known to be involved in SCZ (El-Hadidy, Abdeen, Abd El-Aziz, & Al-Harrass, 2014; Liu et al., 2020; Peerbooms et al., 2011). A large-scale meta-analysis comprising 2,159 EOS and 6,561 HC found a significant variant, i.e., rs1801133, which is located in the fifth exon of *MTHFR* (Guo et al., 2021). Taken together, previous GWAS findings (on common variants) and our rare-variant findings both supported the convergent risk of *MTHFR* in EOS. *STOX1* (Storkhead Box 1) is expressed in the brain and choroid plexus, and is heavily involved in Alzheimer’s disease (AD), especially its late-onset subtype (van Abel, Abdulhamid, Scheper, van Dijk, & Oudejans, 2012; van Abel, Michel, et al., 2012; van Dijk et al., 2010). The expression profiles of *WDHD1* (WD Repeat And HMG-Box DNA Binding Protein 1, highly expressed in the thyrmus) and *ZDHHC14* (Zinc Finger DHHC-Type Palmitoyltransferase 14, with an enhanced expression in plasmacytoid DC) showed immune-related patterns. Specifically, *WDHD1* is correlated with other traits in the immunological domain. Together, the gene-based and gene-set-based association analyses consistently suggested that the immune system and the neural system are involved in SCZ, and perturbations in these two systems may influence neurodevelopment and SCZ onset.

Several limitations should be borne in mind. First, the sample size was modest and insufficient to detect single variants. Second, we did not use a family study design, and therefore could not differentiate between “de novo” and inherited rare-variants. Having said that, we applied the collapsing method, and evaluated the aggregated effect of rare damaging variants in genes and gene-sets. Third, some SCZ participants had missing information of AAO. Moreover, we used a crude method to ascertain AAO in SCZ sample. Lastly, because of the small number of HC, we included unaffected siblings in the control group. Notwithstanding these limitations, our study provided preliminary evidence for the genetic architecture of AAO in SCZ in the Chinese setting. Future research should utilize a large sample of SCZ patients.

To conclude, rare variants contribute to SCZ risk in the Chinese setting. The candidate risk genes for EOS and AOS are related to the neural system, consistent with the neurodevelopmental model of SCZ. On the other hand, candidate risk genes for EOS but not AOS are related to the immune system, suggesting that immune-mediated process may advance the first presentation of SCZ. More research is needed to explore how immune system would modulate SCZ patients’ early neurodevelopment.

## Data Availability

Data produced in the present study are available upon reasonable request to the authors

